# The hospital burden of intergenerational contact with the welfare system: *A whole-of-population liked data study*

**DOI:** 10.1101/2021.11.15.21266370

**Authors:** Alexandra M Procter, Catherine R Chittleborough, Rhiannon M Pilkington, Odette Pearson, Alicia Montgomerie, John W Lynch

## Abstract

**Background:** Intergenerational welfare contact (IWC) is a policy issue because of the personal and social costs of intergenerational disadvantage. We estimated the hospital burden of IWC for children aged 11-20 years.

**Methods:** This linked data study of children born in South Australia, 1991-1995 (n=94,358), and their parent/s (n=143,814) used de-identified data from the Better Evidence Better Outcomes Linked Data platform. Using Australian Government Centrelink data, welfare contact (WC) was defined as parent/s receiving a means-tested welfare payment (low-income, unemployment, disability or caring) when children were aged 11-15, or children receiving payment at ages 16-20. IWC was WC occurring in both parent and child generations. Children were classified as: No WC, parent only WC, child only WC, or IWC. Hospitalisation rates and cumulative incidence were estimated by age and WC group.

**Findings:** IWC affected 34.9% of children, who had the highest hospitalisation rate (133.5 per 1,000 person-years) compared to no WC (46.1 per 1,000 person-years), parent only WC (75.0 per 1,000 person-years), and child only WC (87.6 per 1,000 person-years). Of all IWC children, 43.0% experienced at least one hospitalisation between 11-20, frequently related to injury, mental health, and pregnancy.

**Interpretation:** Children experiencing IWC represent a third of the population aged 11-20. Compared to children with parent-only WC, IWC children had 78% higher hospitalisation rates from age 11 to 20, accounting for over half of all hospitalisations in this age group. Frequent IWC hospitalisation causes were injuries, mental health, and pregnancy.

**Funding:** Medical Research Future Fund, National Health and Medical Research Council, Westpac Scholars Trust.

## INTRODUCTION

In 1848, Engels^1^ questioned “*How is it possible, under such conditions, for the lower class to be healthy and long lived?”*. While there have been major population health advances since the 19^th^ century, health gaps between rich and poor remain stubbornly entrenched even in modern welfare states.^2^ In 2017-18, 1 in 6 Australian children under the age of 15 were living in poverty - less than half the population median household income.^3^ Growing up in socioeconomic disadvantage negatively affects child development, education, labour market, and health outcomes, over a life course and across generations.^4-6^ Income provides access to resources such as nutrition, housing, and medical care which directly and indirectly affect well-being, and thus income is a major social determinant of health.^4,7,8^ In high and middle-income countries welfare systems have the potential to buffer the effects of low income and poverty. Welfare systems, including Centrelink in Australia, aim to transfer income to support those experiencing low income due to joblessness, disability, retirement, and caring responsibilities.^9,10^ Payments are intended to help overcome the income loss from these circumstances, and in some cases foster financial independence through income support for further education.^9,11^ Over 40% of Australia’s cash transfers are targeted at the lowest income quintile, making it the second highest targeted cash transfer system of all OECD countries. Such a highly targeted welfare system supports the notion that welfare receipt is closely related to ongoing poverty and social exclusion.^12^ There is recent policy concern^13^ regarding families who become “trapped” in a cycle of intergenerational welfare contact (IWC). For government and non-government organisations, the policy discussion has been about breaking these cycles of disadvantage.^13,14^

The idea that reliance on welfare is transmitted from one generation to the next is not unique to Australia^11,15,16^ and has been documented in the United States,^17,18^ Canada,^17^ Nordic countries,^19,20^ and United Kingdom.^21^ Despite considerable research documenting that low income is associated with poor health,^4,7,8,22^ there are few studies that examine IWC and health.^11,23^ One study investigated the mediating role of health (limited to BMI, self-reported mental health and physical exercise) in IWC but did not report health outcomes for IWC children.^23^ Previous studies reporting that welfare contact (WC) is associated with poorer health are limited by quality (self-reported measures,^24,25^ selective sample populations,^23-26^ no longitudinal design^15^), lack of intergenerational perspective,^24,26-30^ and WC definitions that include any payment type such as those considered near universal.^11,25-30^ The policy problem of IWC in Australia has been examined by labour economist Cobb-Clark and colleagues (2017) who defined and measured IWC to better understand how disadvantage is transmitted from parents to children.^15^ Using welfare payment eligibility criteria we build on this to develop our preferred definition of means-tested WC that includes welfare payments which are intended to support individuals maintaining a basic standard of living.

The objective of our study is descriptive.^31^ We use whole-of-population linked administrative data that contains Australian government welfare payments linked to jurisdictional inpatient hospitalisations. To our knowledge, this is the first time federal government welfare payment data has been linked to state hospital data in Australia. This linked data study examined the proportion of almost 100,000 South Australian (SA) children born 1991-1995 who experienced IWC and other WC types; and their cause-specific hospitalisation experience from ages 11-20 years. We measured WC for parents when their children were aged 11-15 years, and for the children themselves receiving payment at ages 16-20 years. These birth cohorts and age ranges were selected to maximise the exposure (WC) and outcome (hospitalisation) windows given the data structure involving multiple birth cohorts (Figure 1). While the data platform used is whole-of-population, we acknowledge that this research has not included governance by Aboriginal and Torres Strait Islander people and therefore we chose not to include data of Aboriginal children and families.^32^

## METHODS

### Data sources

The data used for this study were part of the BEBOLD (Better Evidence Better Outcomes through Linked Data) platform which is a whole-of-population data resource including health, education, housing, and welfare systems administrative data.^33^ We used inpatient hospitalisations (Integrated SA Activity Collection), SA Perinatal Statistics Collection, the SA Births, Death and Marriages Registry, the family file (linking parents to children through birth registrations), and Centrelink data. Probabilistic linkage was performed by SA-NT DataLink, an independent third-party linkage agency.^34^ Australian data linkage systems estimate false linkage rates from 0.1%^35^ to 0.5%.^36^ Centrelink data (Data Over Multiple INdividual Occurrences, DOMINO^37^) were linked as part of the BEBOLD Project by the Australian Institute of Health and Welfare (AIHW). DOMINO contains de-identified individual event-based data to provide a longitudinal picture of individual welfare payments by the Commonwealth Department of Social Services.^37^

### Ethics

Ethics approval for use of this data was granted from the University of Adelaide (H-185-2011), the SA Department of Health and Wellbeing (HREC/13/SAH/106), and the AIHW Ethics Committee (EO2017/4/396). Site-Specific Approval for use of hospital data was given by SA Department of Health and Wellbeing (Central SSA/13/SAH/146) and Women’s and Children’s Health Network (SSA/14/WCHN/21).

### Population

Children in SA born 1991-1995 (n=98,571) were eligible to be included in the study (Figure 2). Individuals were identified as Aboriginal and/or Torres Strait Islander if they had at least one record as Aboriginal and/or Torres Strait Islander across inpatient hospitalisations, SA Perinatal Statistics, Centrelink demographics (DOMINO) and SA Births Registry.^38^ We made a conscious decision, in consultation with Aboriginal and Torres Strait Islander co-author and informed by the principles of Indigenous Data Sovereignty, not include Aboriginal and Torres Strait Islander children (n=4,103, 4.2%) .^32^ A further 110 children were excluded as they did not have a birth registration with at least one parent recorded, and n≤5 parents were excluded as they were unable to be linked due to missing information. Analysis was undertaken on children who were neither Aboriginal nor Torres Strait Islander (n=94,358) and their parents (n=143,814). We defined children’s parent/s using birth registration data, where mother and (if acknowledged) co-parent was recorded. Of the children included in the analysis, n=91,192 (96.6%) had both a mother and co-parent recorded in birth registration data and n=3,166 (3.4%) had one parent recorded.

### Hospitalisations

Inpatient hospitalisation data records age at admission and the International Statistical Classification of Diseases and Related Health Problems, Tenth Revision, Australian Modification (ICD-10-AM) code/s for the admission.^39^ Hospitalisation type by chapter heading of the ICD-10-AM was defined using the primary diagnosis ICD-10-AM code recorded for the admission.^39^ Potentially preventable hospitalisations (PPHs) were calculated under a child-centric definition for admission at ages 11-14 and under the standard Australian definition (best suited to adults) at ages 15-20.^40^ Primary diagnosis codes used to identify paediatric Complex Chronic Conditions (CCCs) were adapted to the ICD-10-AM coding system.^41^

### Welfare contact

WC was defined as having received a Centrelink payment where the payment amount was greater than $0 and the payment type (e.g. Youth Allowance, Newstart etc.) was recorded. Only 0.003% of payments had a missing payment type. Parent WC was defined as parents having WC when children were aged 11-15 years, and child WC as the child themselves having WC when aged 16-20 years (Figure 1). IWC was WC occurring in both parent and child generations. Children were categorised in one of four WC groups: No WC, parent only WC, child only WC and IWC.

We examined WC under two constructs: any WC, and ‘means-tested’ WC that only included payments subject to an income and assets test and aimed to assist those experiencing low income or hardship. Any WC included all payment types including those payments which may be considered ‘near universal’ for families with children (e.g. Family Tax Benefit, Baby Bonus, and Child Care Benefit). Our definition of means-tested WC included: Carer Allowance, Carer Payment, Disability Support Pension (DSP), Parenting payment partnered, Parenting payment single, Newstart Allowance, Newstart Mature Age Allowance, Youth Allowance, Partner allowance, and Wife Pension DSP (wife of a disability support pension). The intent of these payment types is detailed in Supplementary Table 1.

### Analysis

For each birth cohort, we calculated the proportion of children who experienced no WC, parent only WC, child only WC and IWC, under both welfare constructs (any WC and means-tested WC).

All cause age-specific and overall admission rates for children aged 11-25 years were calculated by dividing the number of admissions by person-years. Cumulative incidence was the proportion of children who had experienced a hospital admission from age 11 years up to age 20 years. Hospitalisation admission rates and proportion of children experiencing hospitalisations by ICD-10-AM chapter, PPHs, and CCCs were calculated for ages 11-15 years, when children could have been exposed to parent WC, and 16-20 years, reflecting the age group when children could have experienced their own WC. Hospitalisation outcomes were reported across WC types under the means-tested welfare construct.

### Sensitivity analyses

Given the data structure and the years of observation available, we undertook sensitivity analysis to assess potential misclassification due to the limited observation windows for both parent and child WC under the means-tested welfare construct. For children born in 1991, we examined child WC for an extended period (ages 16-24 years) and compared this to child WC at 16-20 years as used in the main analyses. For children born in 1995 we examined parent WC for an extended period (ages 7-15 years) and compared this to parent WC at 11-15 years as in the main analyses. For children born in both 1991 and 1995 we recreated the four WC groups using the extended period of parent or child WC.

Given family structures can change over time, we calculated the proportion of children whose parent/s were defined by birth registration (recorded at the time of birth) compared to parent/s recorded in the Centrelink relationships file (recorded at time of payment receipt). We defined ‘parents’ in the Centrelink relationships file as parent, grandparent or guardian relationships associated with the child. Gender of Centrelink parent relationship were defined using the gender recorded in the Centrelink demographics file. This sensitivity analysis was undertaken on two birth cohorts: children born 1995 and children born 2002.

Analyses were undertaken using STATA MP Version 16, 64-bit (StataCorp, College Station, TX, USA).

## RESULTS

### Welfare contact

Table 1 shows that using any type of welfare payment nearly half (48.9%, n=46,147) of children in SA born 1991-1995 experienced IWC, and only 9.8% (n=9,269) experienced no WC. Using the means-tested definition, IWC was experienced by a third of children (34.9%, n=32,969), with 39.7% of children experiencing no WC, 10.9% parent only WC, and 14.4% child only WC.

**Table 1:** Welfare contact (WC) for children born 1991-1995, n individuals= 94,358 *Means-tested contains the following payment types: Carer Allowance, Carer Payment, Disability Support Pension, Newstart Allowance, Newstart Mature Age Allowance, Parenting payment partnered, Parenting payment single, Partner allowance. Wife Pension Disability Support Pension (wife of a Disability Support pension), Youth Allowance.

| Welfare construct<br>*Year of birth* | No WC |  | Parent only WC |  | Child only WC |  | Intergenerational WC |  | Total |
| --- | --- | --- | --- | --- | --- | --- | --- | --- | --- |
|  | n | % | n | % | n | % | n | % | n |
| Any payment |  |  |  |  |  |  |  |  |  |
| 1991 | 1,988 | 10.6 | 7,438 | 39.7 | 437 | 2.3 | 8,881 | 47.4 | 18,744 |
| 1992 | 1,911 | 10.0 | 7,536 | 39.4 | 339 | 1.8 | 9,335 | 48.8 | 19,121 |
| 1993 | 1,772 | 9.3 | 7,558 | 39.8 | 294 | 1.5 | 9,388 | 49.4 | 19,012 |
| 1994 | 1,815 | 9.6 | 7,226 | 38.4 | 268 | 1.4 | 9,525 | 50.6 | 18,834 |
| 1995 | 1,783 | 9.6 | 7,584 | 40.7 | 262 | 1.4 | 9,018 | 48.4 | 18,647 |
| 1991-1995 | 9,269 | 9.8 | 37,342 | 39.6 | 1,600 | 1.7 | 46,147 | 48.9 | 94,358 |
| Means-tested* |  |  |  |  |  |  |  |  |  |
| 1991 | 7,470 | 39.9 | 2,231 | 11.9 | 2,517 | 13.4 | 6,526 | 34.8 | 18,744 |
| 1992 | 7,554 | 39.5 | 2,193 | 11.5 | 2,673 | 14.0 | 6,701 | 35.0 | 19,121 |
| 1993 | 7,477 | 39.3 | 2,078 | 10.9 | 2,858 | 15.0 | 6,599 | 34.7 | 19,012 |
| 1994 | 7,397 | 39.3 | 1,845 | 9.8 | 2,835 | 15.1 | 6,757 | 35.9 | 18,834 |
| 1995 | 7,609 | 40.8 | 1,913 | 10.3 | 2,739 | 14.7 | 6,386 | 34.2 | 18,647 |
| 1991-1995 | 37,507 | 39.7 | 10,260 | 10.9 | 13,622 | 14.4 | 32,969 | 34.9 | 94,358 |

### All cause hospitalisations

From the perspective of health system burden as ‘incidents’ of hospitalisation, Table 2 shows the all-cause hospitalisation rate among 11-25 year olds was 46.1 per 1,000 person-years among those with no WC and 133.5 per 1,000 person-years for children with IWC. For every age, IWC children had higher admission rates compared to children experiencing no WC, or WC in only one generation, with the exception of age 11 when IWC and parent only WC had similar hospitalisation rates.

**Table 2:** All cause hospital admission rates per 1,000 person-years (py) for children born 1991-1995 by welfare contact (WC), n individuals=94,358; n admissions=105,250

|  |  | No WC |  |  | Parent only WC |  |  | Child only WC |  |  | Intergenerational WC |  |  | Overall |  |  |
| --- | --- | --- | --- | --- | --- | --- | --- | --- | --- | --- | --- | --- | --- | --- | --- | --- |
|  |  | py | n | rate<br>per<br>1,000<br>py | py | n | rate<br>per<br>1,000<br>py | py | n | rate<br>per<br>1,000<br>py | py | n | rate<br>per<br>1,000<br>py | py | n | rate<br>per<br>1,000<br>py |
| Birth cohorts | Age years |  |  |  |  |  |  |  |  |  |  |  |  |  |  |  |
| 1991-1995 | 11 | 37,507 | 1,066 | 28.4 | 10,260 | 692 | 67.4 | 13,622 | 510 | 37.4 | 32,969 | 2,161 | 65.5 | 94,358 | 4,429 | 46.9 |
| 1991-1995 | 12 | 37,507 | 1,187 | 31.6 | 10,260 | 615 | 59.9 | 13,622 | 613 | 45.0 | 32,969 | 2,184 | 66.2 | 94,358 | 4,599 | 48.7 |
| 1991-1995 | 13 | 37,507 | 1,275 | 34.0 | 10,260 | 664 | 64.7 | 13,622 | 624 | 45.8 | 32,969 | 2,335 | 70.8 | 94,358 | 4,898 | 51.9 |
| 1991-1995 | 14 | 37,507 | 1,377 | 36.7 | 10,260 | 766 | 74.7 | 13,622 | 687 | 50.4 | 32,969 | 2,845 | 86.3 | 94,358 | 5,675 | 60.1 |
| 1991-1995 | 15 | 37,507 | 1,496 | 39.9 | 10,260 | 766 | 74.7 | 13,622 | 855 | 62.8 | 32,969 | 3,401 | 103.2 | 94,358 | 6,518 | 69.1 |
| 1991-1995 | 16 | 37,507 | 1,669 | 44.5 | 10,260 | 732 | 71.3 | 13,622 | 1,275 | 93.6 | 32,969 | 4,275 | 129.7 | 94,358 | 7,951 | 84.3 |
| 1991-1995 | 17 | 37,507 | 1,892 | 50.4 | 10,260 | 874 | 85.2 | 13,622 | 1,339 | 98.3 | 32,969 | 4,842 | 146.9 | 94,358 | 8,947 | 94.8 |
| 1991-1995 | 18 | 37,507 | 1,969 | 52.5 | 10,260 | 707 | 68.9 | 13,622 | 1,463 | 107.4 | 32,969 | 5,242 | 159.0 | 94,358 | 9,381 | 99.4 |
| 1991-1995 | 19 | 37,507 | 1,954 | 52.1 | 10,260 | 724 | 70.6 | 13,622 | 1,546 | 113.5 | 32,969 | 5,637 | 171.0 | 94,358 | 9,861 | 104.5 |
| 1991-1995 | 20 | 37,507 | 1,974 | 52.6 | 10,260 | 771 | 75.1 | 13,622 | 1,667 | 122.4 | 32,969 | 5,691 | 172.6 | 94,358 | 10,103 | 107.1 |
| 1991-1995 | 21 | 37,507 | 2,130 | 56.8 | 10,260 | 851 | 82.9 | 13,622 | 1,747 | 128.2 | 32,969 | 5,568 | 168.9 | 94,358 | 10,296 | 109.1 |
| 1991-1994 | 22 | 29,898 | 1,650 | 55.2 | 8,347 | 730 | 87.5 | 10,883 | 1,268 | 116.5 | 26,583 | 4,917 | 185.0 | 75,711 | 8,565 | 113.1 |
| 1991-1993 | 23 | 22,501 | 1,372 | 61.0 | 6,502 | 643 | 98.9 | 8,048 | 952 | 118.3 | 19,826 | 3,903 | 196.9 | 56,877 | 6,870 | 120.8 |
| 1991-1992 | 24 | 15,024 | 916 | 61.0 | 4,424 | 360 | 81.4 | 5,190 | 621 | 119.7 | 13,227 | 2,774 | 209.7 | 37,865 | 4,671 | 123.4 |
| 1991 | 25 | 7,470 | 523 | 70.0 | 2,231 | 189 | 84.7 | 2,517 | 286 | 113.6 | 6,526 | 1,488 | 228.0 | 18,744 | 2,486 | 132.6 |
| Total | 11-25 | 487,470 | 22,450 | 46.1 | 134,364 | 10,084 | 75.0 | 176,480 | 15,453 | 87.6 | 428,821 | 57,263 | 133.5 | 1,227,135 | 105,250 | 85.8 |

From the perspective of a cohort of children experiencing hospitalisation, Table 3 shows that the cumulative ‘incidence’ (proportion of children hospitalised at least once) from ages 11 to 20 years was highest for IWC children (43.0%) compared to children experiencing no WC (23.7%), parent only WC (29.3%), or child only WC (34.3%).

**Table 3:** Cumulative incidence of all cause hospitalisation by welfare contact (WC) and age, n individuals=94,358

| Age<br>years | No WC<br>n individuals=37,507 |  | Parent only WC<br>n individuals=10,260 |  | Child only WC<br>n individuals =13,622 |  | Intergenerational WC<br>n individuals =32,969 |  | Overall<br>n individuals =94,358 |  |
| --- | --- | --- | --- | --- | --- | --- | --- | --- | --- | --- |
|  | n | % | n | % | n | % | n | % | n | % |
| 11 | 875 | **2.3** | 367 | **3.6** | 417 | **3.1** | 1,598 | **4.8** | 3,257 | **3.5** |
| 12 | 1,734 | **4.6** | 671 | **6.5** | 847 | **6.2** | 2,853 | **8.7** | 6,105 | **6.5** |
| 13 | 2,528 | **6.7** | 978 | **9.5** | 1,216 | **8.9** | 4,069 | **12.3** | 8,791 | **9.3** |
| 14 | 3,404 | **9.1** | 1,263 | **12.3** | 1,610 | **11.8** | 5,404 | **16.4** | 11,681 | **12.4** |
| 15 | 4,301 | **11.5** | 1,585 | **15.4** | 2,066 | **15.2** | 6,891 | **20.9** | 14,843 | **15.7** |
| 16 | 5,219 | **13.9** | 1,898 | **18.5** | 2,577 | **18.9** | 8,461 | **25.7** | 18,155 | **19.2** |
| 17 | 6,190 | **16.5** | 2,212 | **21.6** | 3,151 | **23.1** | 9,987 | **30.3** | 21,540 | **22.8** |
| 18 | 7,128 | **19.0** | 2,506 | **24.4** | 3,728 | **27.4** | 11,526 | **35.0** | 24,888 | **26.4** |
| 19 | 8,003 | **21.3** | 2,773 | **27.0** | 4,248 | **31.2** | 12,888 | **39.1** | 27,912 | **29.6** |
| 20 | 8,873 | **23.7** | 3,009 | **29.3** | 4,679 | **34.3** | 14,190 | **43.0** | 30,751 | **32.6** |

### Cause-specific hospitalisations

The proportion of individuals with at least one hospitalisation by condition type (ICD-10-AM chapter heading, PPHs, and CCCs) and WC group is shown for hospitalisations at ages 11-15 years (Table 4) and 16-20 years (Table 5). The most common causes of admission were injuries, poisoning and external causes regardless of WC or age group. A higher proportion of IWC children experienced at least one hospitalisation related to respiratory conditions, mental health, digestive conditions, and pregnancy compared to any other type of WC for both age groups. For each of these causes the proportion was higher at ages 16-20 compared to 11-15 years (e.g. *‘Mental and Behavioural disorders (F00-F09)’* age 11-15 years: 1.4%; age 16-20 years: 3.7%). IWC children had the highest proportion of PPHs compared to children experiencing all other WC types (age 11-15 years: 4.5%; age 16-20 years: 4.8%). Only a small proportion of IWC children experienced CCCs across both age groups (age 11-15 years: 1.9%; age 16-20 years: 2.2%). Similar patterns were observed when taking a health system perspective by reporting the cause-specific (ICD-10-AM chapter heading, PPHs and CCCs) admission rates per 1,000 person-years by WC and age group (Supplementary Tables 2 and 3).

**Table 4:** Proportion of individuals with at least one hospitalisation by condition type and welfare contact (WC) for children born 1991-1995, aged 11-15 years, n individuals=94,358

| Condition type (respective ICD-10-AM codes) | No WC<br>n individuals=37,507 |  | Parent only WC<br>n individuals=10,260 |  | Child only WC<br>n individuals =13,622 |  | Intergenerational WC<br>n individuals=32,969 |  | Overall<br>n individuals =94,358 |  |
| --- | --- | --- | --- | --- | --- | --- | --- | --- | --- | --- |
|  | n | % | n | % | n | % | n | % | n | % |
| Certain infectious & parasitic diseases (A00-B99) | 283 | **0.8** | 109 | **1.1** | 128 | **0.9** | 425 | **1.3** | 945 | **1.0** |
| Neoplasms (C00-D48) | 92 | **0.2** | 84 | **0.8** | 65 | **0.5** | 229 | **0.7** | 470 | **0.5** |
| Diseases of the blood and blood-forming organs and certain disorders involving the immune mechanism (D50-D89) | 24 | **0.1** | 26 | **0.3** | 9 | **0.1** | 41 | **0.1** | 100 | **0.1** |
| Endocrine, nutritional and metabolic disorders (E00-E89) | 90 | **0.2** | 79 | **0.8** | 41 | **0.3** | 197 | **0.6** | 407 | **0.4** |
| Mental and behavioural disorders (F00-F99) | 151 | **0.4** | 50 | **0.5** | 88 | **0.6** | 457 | **1.4** | 746 | **0.8** |
| Diseases of the nervous system (G00-G99) | 112 | **0.3** | 51 | **0.5** | 52 | **0.4** | 285 | **0.9** | 500 | **0.5** |
| Diseases of the eye and adnexa (H00-H59) | 34 | **0.1** | 8 | **0.1** | 17 | **0.1** | 87 | **0.3** | 146 | **0.2** |
| Disease of the ear and mastoid process (H60-H95) | 71 | **0.2** | 41 | **0.4** | 48 | **0.4** | 216 | **0.7** | 376 | **0.4** |
| Diseases of the circulatory system (I00-I99) | 71 | **0.2** | 34 | **0.3** | 34 | **0.2** | 120 | **0.4** | 259 | **0.3** |
| Diseases of the respiratory system (J00-J99) | 504 | **1.3** | 222 | **2.2** | 318 | **2.3** | 1,171 | **3.6** | 2,215 | **2.3** |
| Diseases of the digestive system (K00-K93) | 680 | **1.8** | 252 | **2.5** | 365 | **2.7** | 1,112 | **3.4** | 2,409 | **2.6** |
| Diseases of the skin and subcutaneous tissue system (L00-L099) | 214 | **0.6** | 77 | **0.8** | 128 | **0.9** | 500 | **1.5** | 919 | **1.0** |
| Diseases of the musculoskeletal system and connective tissue (M00-M99) | 205 | **0.5** | 73 | **0.7** | 106 | **0.8** | 385 | **1.2** | 769 | **0.8** |
| Diseases of the genitourinary system (N00-N99) | 204 | **0.5** | 84 | **0.8** | 98 | **0.7** | 354 | **1.1** | 740 | **0.8** |
| Pregnancy, childbirth and the puerperium (O00-O99) | 7 | **0.0** | 6 | **0.1** | 21 | **0.2** | 145 | **0.4** | 179 | **0.2** |
| Certain conditions originating in the perinatal period (P00-P96) | 0 | **0.0** | 0 | **0.0** | 0 | **0.0** | 0 | **0.0** | 0 | **0.0** |
| Congenital malformations, deformations and chromosomal abnormalities (Q00-Q99) | 87 | **0.2** | 37 | **0.4** | 39 | **0.3** | 205 | **0.6** | 368 | **0.4** |
| Symptoms, signs and abnormal clinical and laboratory findings, not elsewhere classified (R00-R99) | 452 | **1.2** | 161 | **1.6** | 211 | **1.5** | 846 | **2.6** | 1,670 | **1.8** |
| Injury, poisoning and certain other consequences of external causes (S00-T98) | 1,670 | **4.5** | 554 | **5.4** | 710 | **5.2** | 2,174 | **6.6** | 5,108 | **5.4** |
| Factors influencing health status and contact with health services (Z00-Z99) | 208 | **0.6** | 102 | **1.0** | 87 | **0.6** | 328 | **1.0** | 725 | **0.8** |
| Potentially preventable hospitalisations (PPHs) | 726 | **1.9** | 296 | **2.9** | 364 | **2.7** | 1,485 | **4.5** | 2,871 | **3.0** |
| Complex Chronic Conditions (CCCs) | 164 | **0.4** | 208 | **2.0** | 79 | **0.6** | 630 | **1.9** | 1,081 | **1.1** |

*Note: children may experience multiple admissions and therefore may fall into multiple ICD-10-AM chapters*

**Table 5:**
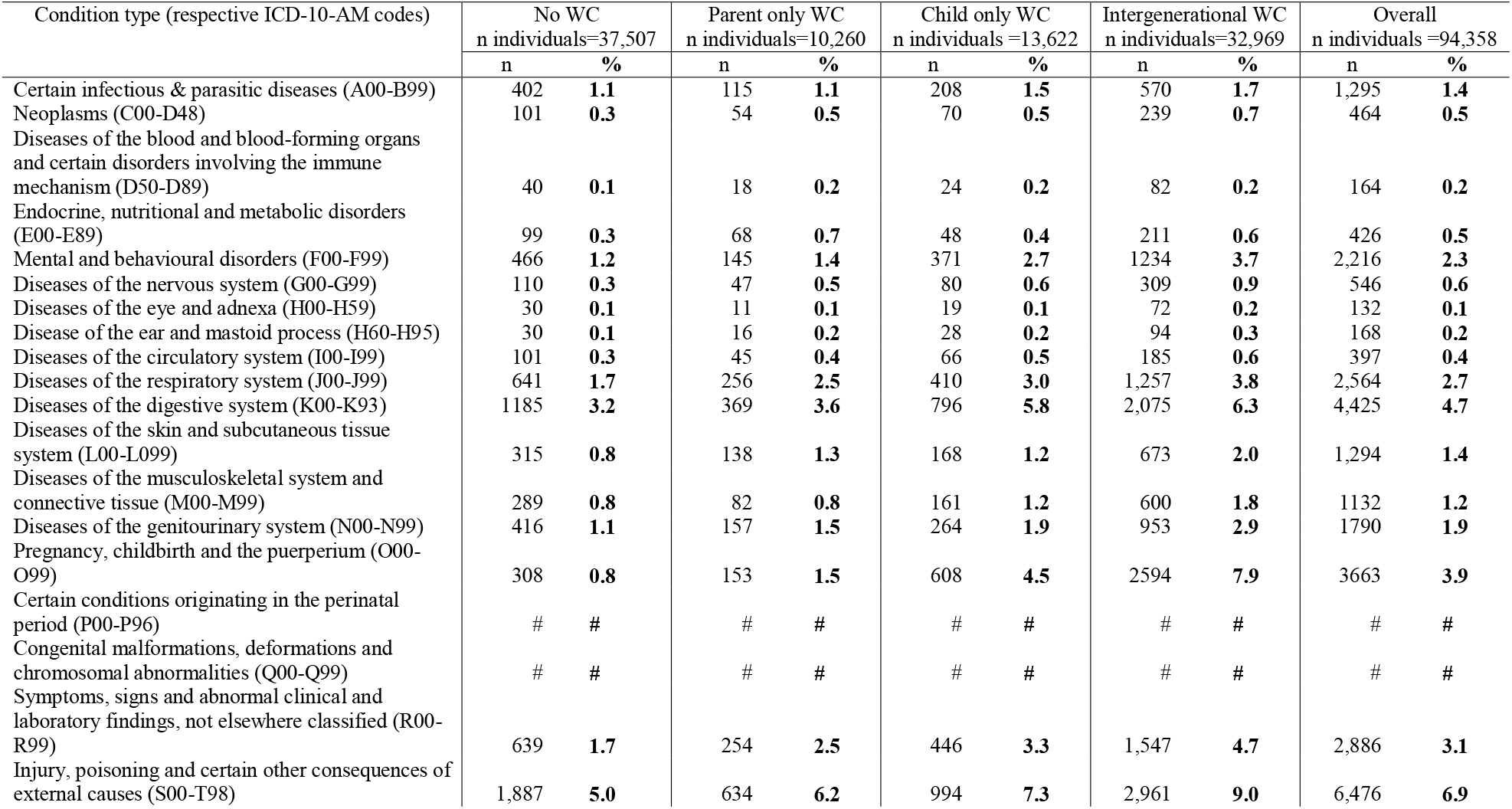
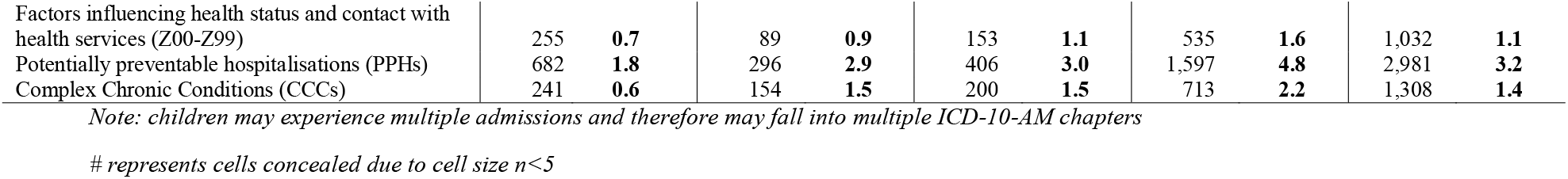
Proportion of individuals with at least one hospitalisation by condition type and welfare contact (WC) for children born 1991-1995, aged 16-20 years, n individuals=94,358

| Condition type (respective ICD-10-AM codes) | No WC<br>n individuals=37,507 |  | Parent only WC<br>n individuals=10,260 |  | Child only WC<br>n individuals =13,622 |  | Intergenerational WC<br>n individuals=32,969 |  | Overall<br>n individuals =94,358 |  |
| --- | --- | --- | --- | --- | --- | --- | --- | --- | --- | --- |
|  | n | % | n | % | n | % | n | % | n | % |
| Certain infectious & parasitic diseases (A00-B99) | 402 | **1.1** | 115 | **1.1** | 208 | **1.5** | 570 | **1.7** | 1,295 | **1.4** |
| Neoplasms (C00-D48) | 101 | **0.3** | 54 | **0.5** | 70 | **0.5** | 239 | **0.7** | 464 | **0.5** |
| Diseases of the blood and blood-forming organs and certain disorders involving the immune mechanism (D50-D89) | 40 | **0.1** | 18 | **0.2** | 24 | **0.2** | 82 | **0.2** | 164 | **0.2** |
| Endocrine, nutritional and metabolic disorders (E00-E89) | 99 | **0.3** | 68 | **0.7** | 48 | **0.4** | 211 | **0.6** | 426 | **0.5** |
| Mental and behavioural disorders (F00-F99) | 466 | **1.2** | 145 | **1.4** | 371 | **2.7** | 1234 | **3.7** | 2,216 | **2.3** |
| Diseases of the nervous system (G00-G99) | 110 | **0.3** | 47 | **0.5** | 80 | **0.6** | 309 | **0.9** | 546 | **0.6** |
| Diseases of the eye and adnexa (H00-H59) | 30 | **0.1** | 11 | **0.1** | 19 | **0.1** | 72 | **0.2** | 132 | **0.1** |
| Disease of the ear and mastoid process (H60-H95) | 30 | **0.1** | 16 | **0.2** | 28 | **0.2** | 94 | **0.3** | 168 | **0.2** |
| Diseases of the circulatory system (I00-I99) | 101 | **0.3** | 45 | **0.4** | 66 | **0.5** | 185 | **0.6** | 397 | **0.4** |
| Diseases of the respiratory system (J00-J99) | 641 | **1.7** | 256 | **2.5** | 410 | **3.0** | 1,257 | **3.8** | 2,564 | **2.7** |
| Diseases of the digestive system (K00-K93) | 1185 | **3.2** | 369 | **3.6** | 796 | **5.8** | 2,075 | **6.3** | 4,425 | **4.7** |
| Diseases of the skin and subcutaneous tissue system (L00-L099) | 315 | **0.8** | 138 | **1.3** | 168 | **1.2** | 673 | **2.0** | 1,294 | **1.4** |
| Diseases of the musculoskeletal system and connective tissue (M00-M99) | 289 | **0.8** | 82 | **0.8** | 161 | **1.2** | 600 | **1.8** | 1132 | **1.2** |
| Diseases of the genitourinary system (N00-N99) | 416 | **1.1** | 157 | **1.5** | 264 | **1.9** | 953 | **2.9** | 1790 | **1.9** |
| Pregnancy, childbirth and the puerperium (O00-O99) | 308 | **0.8** | 153 | **1.5** | 608 | **4.5** | 2594 | **7.9** | 3663 | **3.9** |
| Certain conditions originating in the perinatal period (P00-P96) | # | # | # | # | # | # | # | # | # | # |
| Congenital malformations, deformations and chromosomal abnormalities (Q00-Q99) | # | # | # | # | # | # | # | # | # | # |
| Symptoms, signs and abnormal clinical and laboratory findings, not elsewhere classified (R00-R99) | 639 | **1.7** | 254 | **2.5** | 446 | **3.3** | 1,547 | **4.7** | 2,886 | **3.1** |
| Injury, poisoning and certain other consequences of external causes (S00-T98) | 1,887 | **5.0** | 634 | **6.2** | 994 | **7.3** | 2,961 | **9.0** | 6,476 | **6.9** |
| Factors influencing health status and contact with health services (Z00-Z99) | 255 | **0.7** | 89 | **0.9** | 153 | **1.1** | 535 | **1.6** | 1,032 | **1.1** |
| Potentially preventable hospitalisations (PPHs) | 682 | **1.8** | 296 | **2.9** | 406 | **3.0** | 1,597 | **4.8** | 2,981 | **3.2** |
| Complex Chronic Conditions (CCCs) | 241 | **0.6** | 154 | **1.5** | 200 | **1.5** | 713 | **2.2** | 1,308 | **1.4** |

*Note: children may experience multiple admissions and therefore may fall into multiple ICD-10-AM chapters*
*# represents cells concealed due to cell size $n < 5$*

### Sensitivity analysis

Both sensitivity analyses were consistent with the main findings.

Supplementary Table 4 compares child WC measured at 16-20 years to an extended view measured at 16-24 years for children born in 1991. Under the extended view, an additional n=2,204 children (11.8% of the birth cohort) experienced child only WC and an additional n=677 children (3.6% of birth cohort) experienced IWC. Supplementary Table 5 compares parent WC measured for children born in 1995 when the child was 11-15 years to an extended view measured when the child was 7-15 years. An additional n=912 children (4.9% of the birth cohort) experienced parent only WC and an additional n=659 children (3.5% of the birth cohort) experienced IWC. Increasing observation periods of either parent or child WC increased the portion of the cohort who experiences IWC by ∼3.5%.

Sensitivity analysis was undertaken for children born in 1991 and 1995 to compare parental family structure defined by birth registration data to male and female parents recorded in the Centrelink relationships file. For 96.9% of children who had a female parent recorded in the Centrelink relationships file, that female parent ID matched the mother on the birth registration. For 90% of children with at least one male parent registered in the Centrelink relationships file, that male parent ID matched the co-parent on the birth registration. Only 33% of co-parents in birth registrations were recorded in the Centrelink relationships file as having a relationship to the child.

## DISCUSSION

This study showed that 1 in 3 children (34.9%) born in SA from 1991-1995 experienced IWC defined as means-tested payments designed to support more disadvantaged families raising children, and those children themselves. This is perhaps unsurprising, given cross-sectional survey estimates suggest 1 in 6 Australian children under 15 live in poverty.^3^ Our findings provide a nuanced understanding of IWC as it considered welfare payments with eligibility requirements linked to socioeconomic disadvantage. If we took the same approach as previous research and considered any payment type, 1 in 2 children (48.9%, n=46,147) had experienced IWC. However, this estimate includes near universal payments that normalise the experience of IWC in society. While it is important that such universal supports exist, they have little meaningful interpretation for considering potential interventions^42^ to help break cycles of intergenerational disadvantage which should focus on differences between children with parent only WC and IWC.

We believe this is the first study to report the health burden of IWC using actual welfare payment data. Children with IWC had a 78% higher hospitalisation rate compared to children with parent only WC. We compare IWC children to those experiencing parent only WC as they are the target group we would potentially intervene on to ‘break the cycle’ of IWC.^42^ Of all IWC children, 43% experienced at least one hospitalisation between 11-20 years, most commonly related to injury, respiratory conditions, digestive conditions, mental health, and pregnancy. While welfare receipt has been associated with poorer mental health outcomes,^11,24,26,29^ our study showed IWC children experience even higher mental health hospitalisation rates compared to WC experienced only in one generation. We expected IWC children to have higher injury hospitalisation rates given injury hospitalisation is more frequent in children from the most disadvantaged backgrounds^40,41^ but this had not been quantified before. IWC children having higher hospitalisation rates related to pregnancy and childbirth reflects the inclusion of welfare payments associated with parenting.^15^ A higher proportion of IWC children experienced a PPH which are also associated with socioeconomic disadvantage.^40^ While IWC children were more likely to experience a hospitalisation relating to a CCC, the overall proportion was small (age 11-15 years: 1.9%; age 16-20 years: 2.2%).

This suggests the majority of IWC children are not experiencing ongoing chronic conditions and are likely to be receiving welfare payments due to low income and other indicators of socioeconomic disadvantage rather than chronic poor health.

Children who had a parent with WC were 2.4 times as likely to experience their own WC compared to children with no parent WC. This finding is consistent with previous work both in Australia^15^ and internationally.^18-20^ Progress in breaking cycles of entrenched disadvantage is slow.^13,43^ We show a third of the population experiences IWC and that IWC children contribute a higher hospital burden compared to children who experience no, parent only or child only WC. This distinction is important. It suggests that breaking the intergenerational cycle of WC is important for reducing hospital burden.

### Aboriginal and Torres Strait Islander people and their data

Aboriginal and Torres Strait Islander children and families are over-represented in the welfare system.^9^ Typically, the outcomes for Aboriginal and Torres Strait Islander children and families are compared to the outcomes of non-Indigenous children and families. This is done without any or very little consideration of the context of the phenomena of interest. The Australian welfare system was constructed to improve wellbeing but the conceptual model it was built on was not created for Aboriginal and Torres Strait Islander people. If we were to take an Aboriginal and Torres Strait Islander world view, we would need to use a different conceptual model of what ‘welfare’ means. Such a model would consider family and kinship structure, the sharing of resources, and understand that money does not necessarily have the same meaning, nor reflects the holistic concept of wellbeing, for Aboriginal and Torres Strait Islander families and communities.^44^ A collective decision was made after consultation among the author group, one of whom is an Aboriginal and Torres Strait Islander researcher, to apply the principles of Indigenous Data Sovereignty to control the Aboriginal and Torres Strait Islander data in the BEBOLD data platform. In this case, the South Australian Aboriginal community were not involved in informing this research and so their data were not included. Globally, Indigenous populations are enacting Indigenous Data Sovereignty which is, *‘the rights of Indigenous Peoples to determine the means of collection, access, analysis, interpretation, management, dissemination and re-use of data pertaining to the Indigenous Peoples from whom it has been derived, or to whom it relates’*.^32^ These principles are fundamental and should be considered and applied by all researchers.

### Limitations

We defined WC as a parent or child having received an eligible payment. Consideration of the amount and duration of payment are topics for future research. However, while it may be possible that individuals only received payments for a short period of time this is unlikely given that payments included under the means-tested construct are intended for longer-term income support. Cobb-Clark and colleagues found the likelihood of children experiencing IWC was more closely related to whether the parent received a welfare payment at all, rather than the amount, type or intensity.^15^

Our study defined family structure at the time of childbirth using SA birth registrations, which contains a record of mother and co-parent (if acknowledged). Centrelink records contain additional information regarding relationship that can imply family structure (i.e. parent, grandparent, sibling etc.). We found a high proportion of mothers (96.9%) and co-parents (90%) from birth registration matched the Centrelink relationships file where recorded. Only 33% of birth registration co-parents were recorded in the Centrelink relationships file. If we were to define family structure using the Centrelink relationships file it would not be possible to know which parent relationship reflected the primary caregiver, and our view of two parent families would be substantially limited.

Our estimates of WC type and IWC were limited by the data available. Sensitivity analysis showed that by increasing observation periods of either parent or child WC, the proportion of the cohort who experienced IWC increased by ∼3.5%. While there is no reason to suspect that the association between WC and hospitalisations would be different if we could examine the entire life course of the children, our estimates of IWC should be considered an underestimation.

Private hospital data are not available for linkage in SA but public hospitals account for 92% of admissions classified as an emergency and the only children’s hospital in SA is a public hospital,^45^ suggesting that we have captured the majority of hospitalisations in our analysis. The exclusion of private hospital data may mean we under-estimated cumulative incidence, but we would not expect the associations between WC and hospital burden to differ between children attending public and private hospitals.

## CONCLUSIONS

Our study examined the health burden of IWC using a definition of means-tested payments designed to support disadvantaged families and their children. Children with IWC had a 78% higher hospitalisation rate compared to children with parent only WC, and 190% higher than no WC. The hospital burden associated with IWC children is large, with 43% IWC children experiencing at least one hospitalisation between ages 11-20 years. Means-tested IWC children comprised one-third of the study population, but accounted for over half of all hospital admissions from ages 11-20. Common causes of hospitalisations included PPHs, injuries, mental health, pregnancy and childbirth.

## Data Availability

All data analysed in the present study are not available.

