## Supplementary tables for "The hospital burden of intergenerational contact with the welfare system: *A whole-of-population liked data study*"

**Supplementary table 1:** Welfare payment types included under the means-tested definition

| **Payment type** | **Intention** | **Means testing** | **Activity tested/ mutual obligations** |
|---|---|---|---|
| Carer Allowance | Income supplement paid to an individual who provides daily care and attention at home to a person with disability/illness. | Yes | No |
| Carer Payment | Paid to an individual who is providing constant care for person(s) with disability/illness. | Yes | No |
| Disability Support Pension (DSP) | Individual diagnosed with permanent physical, intellectual, or psychiatric impairment that meets specific manifest eligibility criteria (e.g. permanently blind) | Yes | No |
| | Individual diagnosed with permanent physical, intellectual, or psychiatric impairment which meets minimum impairment threshold AND cannot work for ≥15h/week for 2 years. | | Potentially-dependent on level of impairment due to disability. |
| Parenting payment partnered | Principal carer of ≥1 child who is ≤6 years old | Yes | Yes |
| Parenting payment single | Principal carer of ≥1 child who is ≤8 years old | Yes | Yes |
| Newstart Allowance | Unemployed, looking for work, and willing to work | Yes | Yes |
| Newstart Mature Age Allowance | Unemployed, looking for work, and willing to work, aged 55 years and over | Yes | Yes |
| Youth Allowance | Looking for employment or doing approved activities (e.g. full-time student or Australian apprentice) | Yes | Yes |
| Partner allowance | Member of a couple where partner received qualifying pension (e.g. Newstart, DSP, carer allowance etc.) and recipient has no recent workforce experience. | Yes | Yes |
| Wife Pension DSP (wife of a disability support pension) | Income support payment for female partners of people receiving Disability support pension. | Yes | No |

**Supplementary table 2:** Cause specific admission rates per 1,000 person-years (py) for children born 1991-1995, aged 11-15 years by welfare contact (WC), n individuals=94,358

| Condition type (respective ICD-10-AM codes) | No WC py=187,535 | | Parent only WC py=51,300 | | Child only WC py=68,110 | | Intergenerational WC py=164,845 | | Overall py=471,790 | |
|---|---|---|---|---|---|---|---|---|---|---|
| | n | **Rate per 1,000 py** | n | **Rate per 1,000 py** | n | **Rate per 1,000 py** | n | **Rate per 1,000 py** | n | **Rate per 1,000 py** |
| Certain infectious & parasitic diseases (A00-B99) | 307 | **1.6** | 154 | **3.0** | 135 | **2.0** | 454 | **2.8** | 1,050 | **2.2** |
| Neoplasms (C00-D48) | 101 | **0.5** | 322 | **6.3** | 73 | **1.1** | 499 | **3.0** | 995 | **2.1** |
| Diseases of the blood and blood-forming organs and certain disorders involving the immune mechanism (D50-D89) | 41 | **0.2** | 212 | **4.1** | 12 | **0.2** | 229 | **1.4** | 494 | **1.0** |
| Endocrine, nutritional and metabolic disorders (E00-E89) | 159 | **0.8** | 428 | **8.3** | 71 | **1.0** | 643 | **3.9** | 1,301 | **2.8** |
| Mental and behavioural disorders (F00-F99) | 207 | **1.1** | 63 | **1.2** | 127 | **1.9** | 702 | **4.3** | 1,099 | **2.3** |
| Diseases of the nervous system (G00-G99) | 126 | **0.7** | 73 | **1.4** | 67 | **1.0** | 442 | **2.7** | 708 | **1.5** |
| Diseases of the eye and adnexa (H00-H59) | 39 | **0.2** | 8 | **0.2** | 22 | **0.3** | 114 | **0.7** | 183 | **0.4** |
| Disease of the ear and mastoid process (H60-H95) | 87 | **0.5** | 48 | **0.9** | 66 | **1.0** | 285 | **1.7** | 486 | **1.0** |
| Diseases of the circulatory system (I00-I99) | 86 | **0.5** | 42 | **0.8** | 39 | **0.6** | 137 | **0.8** | 304 | **0.6** |
| Diseases of the respiratory system (J00-J99) | 610 | **3.3** | 296 | **5.8** | 395 | **5.8** | 1,636 | **9.9** | 2,937 | **6.2** |
| Diseases of the digestive system (K00-K93) | 854 | **4.6** | 345 | **6.7** | 450 | **6.6** | 1,430 | **8.7** | 3,079 | **6.5** |
| Diseases of the skin and subcutaneous tissue system (L00-L099) | 289 | **1.5** | 96 | **1.9** | 166 | **2.4** | 625 | **3.8** | 1,176 | **2.5** |
| Diseases of the musculoskeletal system and connective tissue (M00-M99) | 236 | **1.3** | 110 | **2.1** | 116 | **1.7** | 493 | **3.0** | 955 | **2.0** |
| Diseases of the genitourinary system (N00-N99) | 231 | **1.2** | 106 | **2.1** | 110 | **1.6** | 412 | **2.5** | 859 | **1.8** |
| Pregnancy, childbirth and the puerperium (O00-O99) | 7 | **0.0** | 6 | **0.1** | 29 | **0.4** | 182 | **1.1** | 224 | **0.5** |
| Certain conditions originating in the perinatal period (P00-P96) | 0 | **0.0** | 0 | **0.0** | 0 | **0.0** | 0 | **0.0** | 0 | **0.0** |
| Congenital malformations, deformations and chromosomal abnormalities (Q00-Q99) | 96 | **0.5** | 50 | **1.0** | 45 | **0.7** | 266 | **1.6** | 457 | **1.0** |
| Symptoms, signs and abnormal clinical and laboratory findings, not elsewhere classified (R00-R99) | 518 | **2.8** | 200 | **3.9** | 258 | **3.8** | 1,054 | **6.4** | 2,030 | **4.3** |
| Injury, poisoning and certain other consequences of external causes (S00-T98) | 1,977 | **10.5** | 672 | **13.1** | 912 | **13.4** | 2,787 | **16.9** | 6,348 | **13.5** |
| Factors influencing health status and contact with health services (Z00-Z99) | 430 | **2.3** | 272 | **5.3** | 196 | **2.9** | 536 | **3.3** | 1,434 | **3.0** |

| | | | | | | | | | | |
|---|---|---|---|---|---|---|---|---|---|---|
| Potentially preventable hospitalisations (PPHs) | 832 | **4.4** | 341 | **6.6** | 409 | **6.0** | 1,790 | **10.9** | 3,372 | **7.1** |
| Complex Chronic Conditions (CCCs) | 248 | **1.3** | 1,039 | **20.3** | 119 | **1.7** | 1,349 | **8.2** | 2,754 | **5.8** |
| Total | 6,401 | **34.1** | 3,503 | **68.3** | 3,289 | **48.3** | 12,926 | **78.4** | 26,119 | **55.4** |

**Supplementary table 3:** Cause specific admission rates per 1,000 person-years (py) for children born 1991-1995, aged 16-20 years by welfare contact (WC), n individuals =94,358

| Condition type (respective ICD-10-AM codes) | No WC py=187,535 | | Parent only WC py=51,300 | | Child only WC py=68,110 | | Intergenerational WC py=164,845 | | Overall py=471,790 | |
|---|---|---|---|---|---|---|---|---|---|---|
| | n | **Rate per 1,000 py** | n | **Rate per 1,000 py** | n | **Rate per 1,000 py** | n | **Rate per 1,000 py** | n | **Rate per 1,000 py** |
| Certain infectious & parasitic diseases (A00-B99) | 440 | **2.3** | 124 | **2.4** | 222 | **3.3** | 654 | **4.0** | 1,440 | **3.1** |
| Neoplasms (C00-D48) | 224 | **1.2** | 149 | **2.9** | 150 | **2.2** | 525 | **3.2** | 1,048 | **2.2** |
| Diseases of the blood and blood-forming organs and certain disorders involving the immune mechanism (D50-D89) | 77 | **0.4** | 134 | **2.6** | 46 | **0.7** | 398 | **2.4** | 655 | **1.4** |
| Endocrine, nutritional and metabolic disorders (E00-E89) | 160 | **0.9** | 139 | **2.7** | 99 | **1.5** | 700 | **4.2** | 1,098 | **2.3** |
| Mental and behavioural disorders (F00-F99) | 665 | **3.5** | 188 | **3.7** | 817 | **12.0** | 2,372 | **14.4** | 4,042 | **8.6** |
| Diseases of the nervous system (G00-G99) | 145 | **0.8** | 68 | **1.3** | 134 | **2.0** | 534 | **3.2** | 881 | **1.9** |
| Diseases of the eye and adnexa (H00-H59) | 35 | **0.2** | 12 | **0.2** | 35 | **0.5** | # | **#** | 171 | **0.4** |
| Disease of the ear and mastoid process (H60-H95) | 33 | **0.2** | 19 | **0.4** | 37 | **0.5** | 135 | **0.8** | 224 | **0.5** |
| Diseases of the circulatory system (I00-I99) | 124 | **0.7** | 75 | **1.5** | 90 | **1.3** | 242 | **1.5** | 531 | **1.1** |
| Diseases of the respiratory system (J00-J99) | 769 | **4.1** | 309 | **6.0** | 498 | **7.3** | 1,726 | **10.5** | 3,302 | **7.0** |
| Diseases of the digestive system (K00-K93) | 1,374 | **7.3** | 481 | **9.4** | 956 | **14.0** | 2,536 | **15.4** | 5,347 | **11.3** |
| Diseases of the skin and subcutaneous tissue system (L00-L099) | 513 | **2.7** | 181 | **3.5** | 245 | **3.6** | 936 | **5.7** | 1,875 | **4.0** |
| Diseases of the musculoskeletal system and connective tissue (M00-M99) | 359 | **1.9** | 112 | **2.2** | 219 | **3.2** | 755 | **4.6** | 1,445 | **3.1** |
| Diseases of the genitourinary system (N00-N99) | 495 | **2.6** | 188 | **3.7** | 363 | **5.3** | 1,203 | **7.3** | 2,249 | **4.8** |
| Pregnancy, childbirth and the puerperium (O00-O99) | 393 | **2.1** | 215 | **4.2** | 1,009 | **14.8** | 4,918 | **29.8** | 6,535 | **13.9** |
| Certain conditions originating in the perinatal period (P00-P96) | # | **#** | # | **#** | # | **#** | # | **#** | # | **#** |
| Congenital malformations, deformations and chromosomal abnormalities (Q00-Q99) | # | **#** | # | **#** | # | **#** | # | **#** | # | **#** |
| Symptoms, signs and abnormal clinical and laboratory findings, not elsewhere classified (R00-R99) | 727 | **3.9** | 301 | **5.9** | 568 | **8.3** | 2,015 | **12.2** | 3,611 | **7.7** |
| Injury, poisoning and certain other consequences of external causes (S00-T98) | 2,362 | **12.6** | 789 | **15.4** | 1,333 | **19.6** | 4,068 | **24.7** | 8,552 | **18.1** |

| | | | | | | | | | | |
|---|---|---|---|---|---|---|---|---|---|---|
| Factors influencing health status and contact with health services (Z00-Z99) | 493 | **2.6** | 292 | **5.7** | 444 | **6.5** | 1,697 | **10.3** | 2,926 | **6.2** |
| Potentially preventable hospitalisations (PPHs) | 754 | **4.0** | 361 | **7.0** | 471 | **6.9** | 2,041 | **12.4** | 3,627 | **7.7** |
| Complex Chronic Conditions (CCCs) | 4,171 | **8.8** | 584 | **3.1** | 579 | **11.3** | 645 | **9.5** | 2,366 | **14.4** |
| Total | 9,458 | **50.4** | 3,808 | **74.2** | 7,290 | **107.0** | 25,687 | **155.8** | 46,243 | **98.0** |

*# represents cells concealed due to cell size n<5*

**Supplementary table 4:** Means-tested welfare contact (WC) groups when child WC is measured at ages 16-20 years or 16-24 years, children born 1991, n individuals=18,744

| WC when child WC is measured at ages 16-20 years | WC when child WC is measured at ages 16-24 years | | | | | | | | | | | | | |
|---|---|---|---|---|---|---|---|---|---|---|---|---|---|---|
| | No WC | | | Parent only WC | | | Child only WC | | | IWC | | | Total | |
| | n | Row% | Col% | n | Row% | Col% | n | Row% | Col% | n | Row% | Col% | n | Col% |
| No WC | 5,266 | 70.5 | 100.0 | 0 | 0.0 | 0.0 | 2,204 | 29.5 | 46.7 | 0 | 0.0 | 0.0 | 7,470 | 39.9 |
| Parent only WC | 0 | 0.0 | 0.0 | 1,554 | 69.7 | 100.0 | 0 | 0.0 | 0.0 | 677 | 30.3 | 9.4 | 2,231 | 11.9 |
| Child only WC | 0 | 0.0 | 0.0 | 0 | 0.0 | 0.0 | 2,517 | 100.0 | 53.3 | 0 | 0.0 | 0.0 | 2,517 | 13.4 |
| IWC | 0 | 0.0 | 0.0 | 0 | 0.0 | 0.0 | 0 | 0.0 | 0.0 | 6,526 | 100.0 | 90.6 | 6,526 | 34.8 |
| Total | 5,266 | 28.1 | 100.0 | 1,554 | 8.3 | 100.0 | 4,721 | 25.2 | 100.0 | 7,203 | 38.4 | 100.0 | 18,744 | 100.0 |

**Supplementary table 5:** Means-tested welfare contact (WC) groups when parent WC is measured at child age 11-15 years and 7-15 years, children born 1995, n individuals=18,647

| WC when parent WC is measured at ages 11-15 years | WC when parent WC is measured at ages 7-15 years | | | | | | | | | | | | | |
|---|---|---|---|---|---|---|---|---|---|---|---|---|---|---|
| | No WC | | | Parent only WC | | | Child only WC | | | IWC | | | Total | |
| | n | Row% | Col% | n | Row% | Col% | n | Row% | Col% | n | Row% | Col% | n | Col% |
| No WC | 6,697 | 88.0 | 100.0 | 912 | 12.0 | 32.3 | 0 | 0.0 | 0.0 | 0 | 0.0 | 0.0 | 7,609 | 40.8 |
| Parent only WC | 0 | 0.0 | 0.0 | 1,913 | 100.0 | 67.7 | 0 | 0.0 | 0.0 | 0 | 0.0 | 0.0 | 1,913 | 10.3 |
| Child only WC | 0 | 0.0 | 0.0 | 0 | 0.0 | 0.0 | 2,080 | 75.9 | 100.0 | 659 | 24.1 | 9.4 | 2,739 | 14.7 |
| IWC | 0 | 0.0 | 0.0 | 0 | 0.0 | 0.0 | 0 | 0.0 | 0.0 | 6,386 | 100.0 | 90.6 | 6,386 | 34.2 |
| Total | 6,697 | 35.9 | 100.0 | 2,825 | 15.1 | 100.0 | 2,080 | 11.2 | 100.0 | 7,045 | 37.8 | 100.0 | 18,647 | 100.0 |
